# Seroprevalence of SARS-CoV-2 antibodies in Saint Petersburg, Russia: a population-based study

**DOI:** 10.1101/2020.11.02.20221309

**Authors:** Anton Barchuk, Dmitriy Skougarevskiy, Kirill Titaev, Daniil Shirokov, Yulia Raskina, Anastasia Novkunkskaya, Petr Talantov, Artur Isaev, Ekaterina Pomerantseva, Svetlana Zhikrivetskaya, Lubov Barabanova, Vadim Volkov

**Affiliations:** European University at St. Petersburg, Shpalernaya Ulitsa, 1, St. Petersburg, Russia, 191187; Clinic “Scandinavia” (LLC Ava-Peter), Ilyushina Ulitsa, 4-1, St. Petersburg, Russia, 197372; The Russian Academy of Sciences Commission for Counteracting the Falsification of Scientific Research, Leingradsky Prospekt, 14, Moscow, Russia, 119991; Center of Genetics and Reproductive Medicine GENETICO LLC, Ulitsa Gubkina, 3-1, Moscow, Russia, 119333; Human Stem Cells Institute, Ulitsa Gubkina, 3-2, Moscow, Russia, 119333; N.N. Petrov National Research Medical Center of Oncology, Pesochny, Leningradskaya Ulitsa, 68, St. Petersburg, Russia, 197758; Health Sciences, Faculty of Social Sciences, Tampere University, Arvo Ylpön katu 34, Tampere, Finland, 33520; ITMO University, Kronverksky Prospekt, 49, St. Petersburg, Russia, 197101

**Keywords:** COVID-19, seroepidemiologic study, SARS-CoV-2 infection antibody testing, selection bias

## Abstract

**Background:** Estimates from initial SARS-CoV-2 serological surveys were likely to be biased due to convenience sampling whereas large-scale population-based serosurveys could be biased due to non-response. This study aims to estimate the seroprevalence of SARS-CoV-2 infection in Saint Petersburg, Russia accounting for non-response bias.

**Methods:** We recruited a random sample of adults residing in St. Petersburg with random digit dialling. Computer-assisted telephone interview was followed by an invitation for an antibody test with randomized rewards for participation. Blood samples collected between May 27, 2020 and June 26, 2020 were assessed for anti-SARS-CoV-2 antibodies using two tests — CMIA and ELISA. The seroprevalence estimates were corrected for non-response bias, test sensitivity, and specificity. Individual characteristics associated with seropositivity were assessed.

**Findings:** 66,250 individuals were contacted, 6,440 adults agreed to be interviewed and were invited to participate in the serosurvey. Blood samples were obtained from 1038 participants. Naïve seroprevalence corrected for test characteristics was 9.0% [95% CI 7.2–10.8] by CMIA and 10.8% [8.8–12.7] by ELISA. Correction for non-response bias decreased seroprevalence estimates to 7.4% [5.7–9.2] for CMIA and to 9.3% [7.4–11.2] for ELISA. The most pronounced decrease in non-response bias-corrected seroprevalence was attributed to the history of any illnesses in the past 3 months and COVID-19 testing. Besides that seroconversion was negatively associated with smoking status, self-reported history of allergies and changes in hand-washing habits.

**Interpretation:** These results suggest that even low estimates of seroprevalence in Europe’s fourth-largest city can be an overestimation in the presence of non-response bias. Serosurvey design should attempt to identify characteristics that are associated both with participation and seropositivity. Further population-based studies are required to explain the lower seroprevalence in smokers and participant reporting allergies.

**Funding:** Polymetal International plc

## Introduction

Serological surveys in the midst of COVID-19 pandemic address the issue of underestimation of the number of cases registered officially with RT-PCR using material from nasopharyngeal swabs [1; 2]. They use blood antibody tests that are markers of past infection. WHO recommends serological surveys to monitor COVID-19 spread [3]. However, estimates from serological surveys can be also biased. Estimates can be distorted by non-response bias, non-representativeness of the study sample, and imperfect test characteristics. Previous serological surveys so far have all but focused on the former [4–10]. This poses a significant problem when some observed factors that influence the decision to participate in the survey may be also associated with test results [11]. Non-response or self-selection bias has been widely acknowledged in descriptive epidemiology [12–15]. In particular, it has been predominantly addressed in seroprevalence surveys of HIV [16].

In this paper we present seroprevalence estimates coming from the first cross-sectional data of our longitudinal study with serial sampling to assess the spread COVID-19 in Saint Petersburg, Russia conducted between May 27 and June 26 2020. St. Petersburg is the second largest city in the country and fourth largest in Europe with the population of approximately 5.2 mln. The first case in the city was registered on 5 March, 2020 and 36,667 cases (7.1 per 1000) were reported as of 31 August, 2020. The study of the spread of COVID-19 in St. Petersburg was established to estimate the extent of epidemic in a population-based manner, and, to the best of our knowledge, this was the first COVID-19 serological survey in the country. Our primary aim was to compare naïve and non-response bias-adjusted seroprevalence to show the utmost importance of rigorous serosurvey designs. We report how various observable characteristics of individuals shift the naïve prevalence estimates when accounted for and carefully address possible sources of bias. Finally, we provide observable characteristics of surveyed individuals that are associated with risk of seroconversion in a population-based study.

## Methods

### Study design and participants

The St. Petersburg COVID-19 study is a population-based epidemiological survey of random sample from the adult population to assess the seroprevalence of anti-SARS-CoV-2 antibodies. The study is conducted as a longitudinal study with serial sampling from the same individuals. The study involved one phone-based survey followed by an individual invitation to the clinic, one paper-based survey, and blood sample collection for antibody testing. Interviews were carried out between May 21, 2020 and June 25, 2020. Blood samples were collected between May 27, 2020 and June 26, 2020.

Eligible individuals were adults residing in St. Petersburg older than 18 years and recruited using the random digit dialling (RDD) method. RDD was accompanied by the computer assisted telephone interviewing (CATI) in order to collect information on both individuals who accepted and declined invitation for testing. Residents of St. Petersburg are almost universal mobile phone users, with 99.5% of households having mobile phones as of 2016 (see Supplementary Appendix Table A3). Participants from six distant districts of the city located too far away from the test site were excluded leaving 12 central districts of the city with population of approximately 4.3 mln. The full study protocol is available online (https://eusp.org/sites/default/files/inline-files/EU_SG-Russian-Covid-Serosurvey-Protocol-CDRU-001_en.pdf).

### Procedures

RDD was carried out using area prefixes of mobile phone numbers to include only mobile phone users in St. Petersburg. The individuals who had answered the call were asked to answer 25 questions on demographics, marital status, education level, income level, past history of illnesses, travelling abroad, household size, social contacts, and visits to public places during lockdown (see full questionnaire in the study protocol). Refusal to participate in blood sampling was also recorded. We have also randomly incentivized respondents to participate in the study by offering complimentary taxi transit to and from the clinic test site for approximately 25% of those who agreed to go through CATI.

Those who had agreed to take part in antibody testing were later contacted by the clinic call center and were assigned an appointment date for blood sampling. The participants signed consent forms and filled out additional paper-based survey forms in the clinic on the day of the visit. Forms included question on the medical history, history of allergies, smoking, alcohol consumption, chronic diseases and medication taken regularly. Blood sampling started on May 27, 2020 and was planned for two weeks but was prolonged till June 26, 2020 because of low participation rates.

### Laboratory tests

We assessed anti-SARS-CoV-2 antibodies using two tests. Serum samples were tested using chemiluminescent microparticle immunoassay (CMIA) Abbott Architect SARS-CoV-2 IgG on the Abbott ARCHITECT® i2000sr platform (Abbott Laboratories, Chicago, USA) that detects immunoglobulin class G (IgG) antibodies to the nucleocapsid protein of SARS-CoV-2 (cutoff for positivity 1.4). In addition to that blood samples were also tested by enzyme-linked immunosorbent assay (ELISA) using CoronaPass total antibodies test (Genetico, Moscow, Russia) that detects total antibodies (cutoff for positivity 1.0) and is based on recombinant receptor binding domain of the spike protein of SARS-CoV-2 (Department of Microbiology, Icahn School of Medicine at Mount Sinai, New York, NY, USA). We simultaneously report seroprevalence based on CMIA and ELISA.

### Sample size

Initial sample size of 1550 participants was calculated assuming prevalence of 20% and test sensitivity (100%) and specificity (99.6%) for our CMIA test with sampling error was 2% using a 95% confidence interval (see Supplementary Appendix Figure A1).[17] After receiving the preliminary results (for 500 individuals), we reduced the sample size by assuming 10% prevalence that gave us a target sample size of 882 participants, that was rounded to 1000 participants.

### Statistical analysis

The primary aim of the study was to assess the seroprevalence of antibodies to SARS-CoV-2 in serum samples based on CMIA tests and ELISA tests accounting for non-response bias and test characteristics (sensitivity and specificity). Seroprevalence was defined as the proportion of those tested positive to all participants. Non-response was assessed by comparison of answers provided during the CATI by those visited the test site and all other surveyed.

To understand the direction of non-response bias in our data we estimated a binomial probit regression of individual agreement to participate in the study and offer his/her blood sample on observable characteristics. We used this fitted model to compute conditional probability to participate in the study (holding all but one variable at mean levels at a time). Our bivariate probit model is formally introduced in Statistical Appendix App. 1).

We analyse variables obtained from CATI and the clinic paper-based survey (ordered or unordered factor variables), and results of antibody tests (binary variables). Participant age was split into groups (18–34, 35–49, 50–64, or ≥65 years old).

In the secondary analyses we also assessed seroprevalence by week based on the date of interview and the date of blood sampling. In subgroup analysis we first compared seroprevalence estimates corrected for non-response between different groups of individuals based on their answers in CATI. To explore individual risk factors for test positivity and obtain prevalence ratios we estimated a generalised linear model with Poisson distribution and a log link restricted to data from participants who completed clinic paper-based survey. We have entertained the possibility to use robust variance-covariance matrix in our adjusted prevalence ratio analysis. However, such adjustment narrowed the confidence intervals rendering our adjusted estimates less conservative [18]. For this reason we report confidence intervals from the unadjusted variance-covariance matrix.

In sensitivity analysis we explored how inclusion of different sets of observable characteristics of individuals (namely, travel history, face mask use, public transport use, visits to public places and others) in the model that corrected seroprevalence for non-response influenced the results. We also applied alternative definitions of seroprevalence (test combination either favouring sensitivity or specificity). To account for possible sample non-representativeness in sensitivity analysis we computed raking weights to match the survey age group and educational attainment proportions in 2016 representative survey of adult city population (see Supplementary Appendix Table A3 for description of this survey and the target proportions). R package anesrake was used to compute the weights [19]. We then estimated seroprevalence on re-weighted data.

We treated refusals to answer certain phone or paper-based survey questions as missing data, for this reason the results onwards are considered after listwise deletion of observations with missing variables.

All reported seroprevalence results were also corrected for test characteristics using the manufacturer’s validation data — sensitivity (100% and 98.7%) and specificity (99.6% and 100%) for CMIA and ELISA test, respectively [20]. Standard errors were computed with delta method. Detailed description of statistical analysis is provided in Statistical Appendix App. 1).

### Data sharing

All analyses were conducted in R with the aid of GJRM package [21], study data and code is available online (https://github.com/eusporg/spb_covid_study20).

### Ethical considerations and study registration

The study was approved by the Research Planning Board of European University at St. Petersburg (on May 20, 2020) and the Ethic Committee of the Clinic “Scandinavia” (on May 26, 2020). The study was registered with the following identifiers: NCT04406038 and ISRCTN11060415.

## Results

### Participation rates

Between May 21 and June 25, 2020 66,250 individuals were reached using RDD. Of 13,071 respondents agreed to participate in the CATI 6,671 were excluded for various reasons (see Figure 1). The resulting 6,400 individuals responded to CATI questionnaire (see Supplementary Appendix Table A2 for details regarding missing records on variables of interest). The respondents were representative of the city population in terms of their gender, employment status, and household size, but were younger than the adult city population as of 2016 and had higher levels of educational attainment (see Supplementary Appendix Table A3).

**Figure 1.**
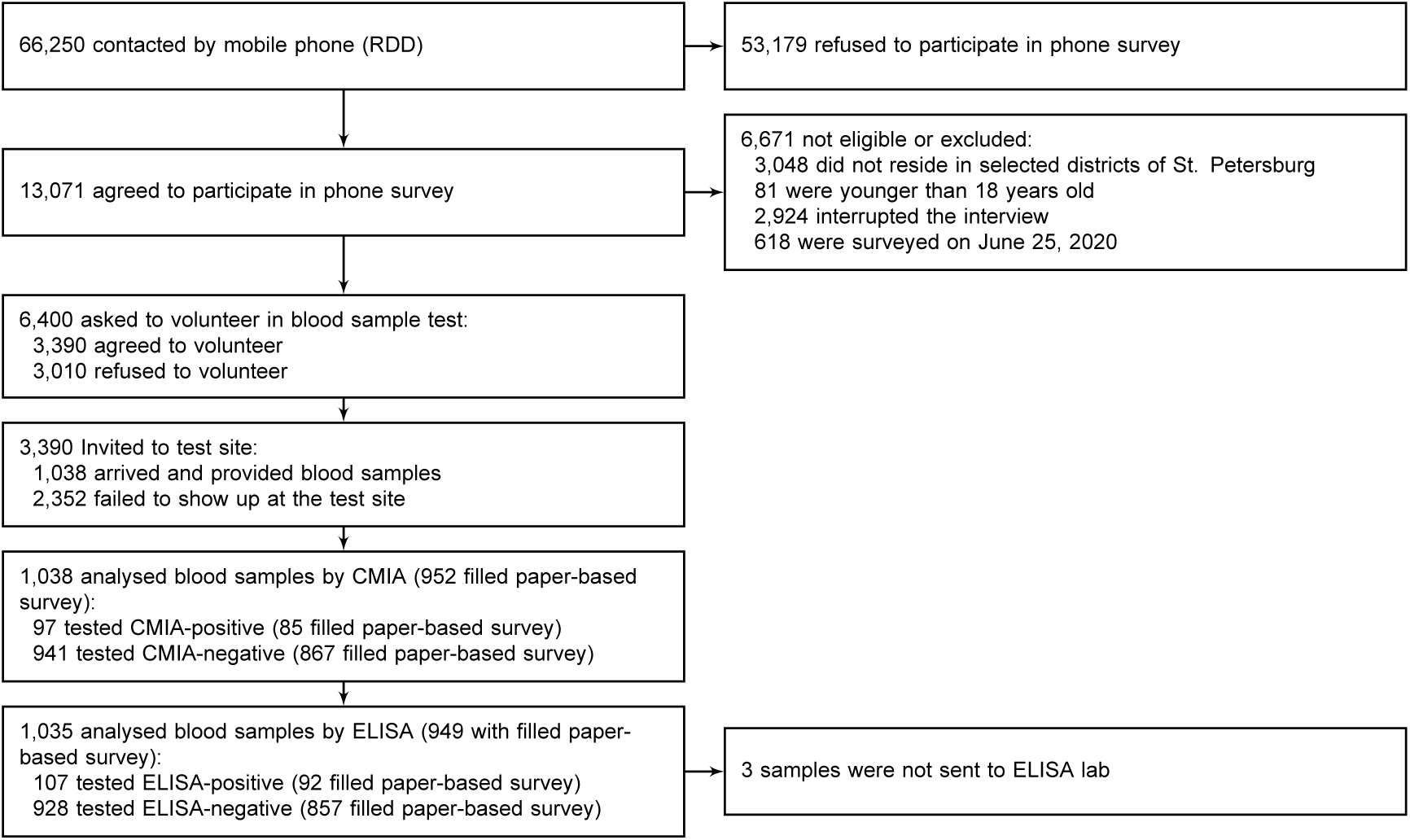
Flow chart of participants’ progress through the St. Petersburg seroprevalence study.

3,390 of surveyed individuals agreed to receive a phone call from the clinic and schedule a visit for antibody testing. Between May 27 and June 26, 2020 only 1038 individuals that satisfied eligibility criteria visited the clinic and provided blood samples (16.2% and 30.6% of those who were interviewed and agreed to participate in serosurvey, respectively). The rest declined the invitation or did not show up at the test site. 1038 CMIA tests and 1035 ELISA tests were eventually performed on eligible individuals. The clinic-visiting participants have also filled out 965 clinic paper-based survey forms.

652 (62.8%) of 1,038 participants were women; 396 (38.2%) were aged 18-34 years, 357 (34.4%) were aged 35-49 years, 218 (21.0%) were aged 50-64 years, and 67 (6.5%) were older than 65 years, the majority of participants lived in multiple-person households, 843 (81.2%) (see Supplementary Appendix Table A2 for summary statistics on phone survey respondents and tested individuals).

In the course of the study we observed the gradual attrition of participants. Compared with the individuals who limited their participation to the CATI, participants who took part in antibody testing were younger, more likely to be female, report a higher education level, experience illnesses in the previous 3 months, report a history of previous COVID-19 testing and a change in their hand-washing habits during the epidemic. Our attempt to randomly incentivize respondents to take part in the study by offering taxi did not reach its purpose. (see Supplementary Appendix Figure A2a).

### Seroprevalence estimates

Between May 27 and June 26, 2020, 115 positive results were reported by any test (97 positive tests out of 1038 were reported by CMIA and 107 positive tests out of 1035 were reported by ELISA). 30 of these 115 (26.1%) individuals with any positive test result did not report any symptoms of past illnesses in the previous 3 months. Naïve seroprevalence corrected for test specificity and sensitivity was 9.0% (95% CI 7.2–10.8) by CMIA and 10.8% (8.8– 12.7) by ELISA (see Table 1). When we accounted for non-response bias with respect to demographic and socioeconomic characteristics our seroprevalence point estimates did not change considerably. Inclusion of characteristics associated with seroprevalence as regressors in our single imputation model shifted point estimates of seroprevalence downwards and after adjustment for all aforementioned characteristics in the model seroprevalence was 7.4% (95% CI 5.7–9.2) for CMIA and to 9.3% (7.4–11.2) for ELISA.

**Table 1.** SARS-CoV-2 seroprevalence estimates from bivariate probit models with different sets of individual characteristics for non-response correction

| Regressors included in bivariate probit model | CMIA |  |  |  | ELISA |  |  |  |
| --- | --- | --- | --- | --- | --- | --- | --- | --- |
|  | Number of participants |  | Seroprevalence (95% CI) |  | Number of participants |  | Seroprevalence (95% CI) |  |
|  | Interviewed | Tested | Naïve | Single imputation | Interviewed | Tested | Naïve | Single imputation |
| Demographic characteristics | 6400 | 1038 | 9.0% (7.2-10.8) | 8.7% (7.0-10.5) | 6397 | 1035 | 10.5% (8.6-12.4) | 10.1% (8.3-12.0) |
| Demographic and socioeconomic characteristics | 6063 | 999 | 9.2% (7.4-11.1) | 9.0% (7.0-11.0) | 6061 | 997 | 10.8% (8.8-12.7) | 10.7% (8.6-12.9) |
| Characteristics associated with seropositivity | 6267 | 1026 | 9.0% (7.2-10.8) | 7.1% (5.6-8.7) | 6264 | 1023 | 10.5% (8.6-12.4) | 8.6% (6.9-10.3) |
| Demographics, socioeconomic status and characteristics associated with seropositivity | 5953 | 990 | 9.2% (7.4-11.1) | 7.4% (5.7-9.2) | 5951 | 988 | 10.8% (8.8-12.7) | 9.1% (7.2-10.9) |

“Demographic characteristics” means the following variables: individual age group (18-34, 35-49, 50-64, 65+ years old) and sex. “Socioeconomic characteristics” means the following variables: higher education status and higher self-reported income level. “Characteristics associated with seropositivity” means the following variables: history of illness in the last 3 months, history of COVID-19 testing, whether respondent lives alone, change in hand washing habits during pandemic, week of the phone interview, and city district. All models include a variable indicating random offer of taxi transportation to and from the clinic test site for interviewed participants. All estimates are corrected for tests characteristics (see Statistical appendix for details).

### Secondary subgroup analysis

Seroprevalence was similar between men and women and was slightly lower in the older (65+) age group (see Table 2). The seroprevalence was higher for individuals who reported past history of illnesses — (15.1% (95% CI 11.6–18.6) for CMIA and 20.0% (95% CI 14.8–25.2) for ELISA) compared to those who did not (3.8% (95% CI 2.1–5.5 for CMIA and 7.4% (95% CI 5.4–9.3 for ELISA). It was also higher for individuals who reported past history of COVID-19 tests, but was slightly lower in individuals who reported that they started washing hands more often since the onset of pandemic and lived alone. There was noticeable variation in seropositivity between city districts (see Figure 2).

**Table 2.** Seroprevalence of SARS-CoV-2 in subgroups of participants

|  |  | CMIA |  |  | ELISA |  |  |
| --- | --- | --- | --- | --- | --- | --- | --- |
|  |  | Number of participants |  | Seroprevalence (95% CI) | Number of participants |  | Seroprevalence (95% CI) |
|  |  | Interviewed | Tested |  | Interviewed | Tested |  |
| Overall |  | 5953 | 990 | 7.4% (5.7-9.2) | 5951 | 988 | 9.1% (7.2-10.9) |
| Age groups | 18-34 | 2228 | 388 | 7.8% (5.2-10.5) | 2227 | 387 | 11.3% (8.1-14.4) |
|  | 35-49 | 1916 | 342 | 6.5% (4.0-9.0) | 1915 | 341 | 7.4% (4.8-10.1) |
|  | 50-64 | 1159 | 199 | 10% (6.0-14.0) | 1159 | 199 | 10.8% (6.6-15.0) |
|  | 65+ | 650 | 61 | 4.1% (0.0-8.8) | 650 | 61 | 3.1% (0.0-7.2) |
| Sex | Female | 3505 | 623 | 7.5% (5.5-9.6) | 3505 | 623 | 8.7% (6.5-10.9) |
|  | Male | 2448 | 367 | 7.3% (4.6-9.9) | 2446 | 365 | 9.5% (6.6-12.5) |
| Higher education | No | 1928 | 169 | 7.4% (3.8-10.9) | 1927 | 168 | 9.7% (5.7-13.7) |
|  | Yes | 4025 | 821 | 7.5% (5.7-9.3) | 4024 | 820 | 8.7% (6.8-10.6) |
| Higher income | No | 3402 | 491 | 6.6% (4.4-8.8) | 3402 | 491 | 8.6% (6.2-11) |
|  | Yes | 2551 | 499 | 8.5% (6.1-11.0) | 2549 | 497 | 9.7% (7.1-12.3) |
| Respondent lives alone | No | 4857 | 805 | 8.0% (6.0-9.9) | 4855 | 803 | 9.8% (7.7-12.0) |
|  | Yes | 1096 | 185 | 5.1% (2.0-8.1) | 1096 | 185 | 5.5% (2.5-8.6) |
| History of illness in the last 3 months | No | 4047 | 548 | 3.8% (2.1-5.5) | 4046 | 547 | 5.0% (3.1-7.0) |
|  | Yes | 1906 | 442 | 15.1% (11.6-18.6) | 1905 | 441 | 17.6% (13.9-21.3) |
| History of COVID-19 testing | No | 5038 | 762 | 5.4% (3.7-7.1) | 5036 | 760 | 7.2% (5.2-9.1) |
|  | Yes | 915 | 228 | 18.6% (13.6-23.6) | 915 | 228 | 19.4% (14.4-24.5) |
| Change in hand washing habits during pandemic | No | 2029 | 279 | 9.6% (6.3-12.9) | 2029 | 279 | 11.8% (8.2-15.4) |
|  | Yes | 3924 | 711 | 6.3% (4.5-8.1) | 3922 | 709 | 7.6% (5.7-9.6) |

All estimates are from the model that includes demographics, socioeconomic status and characteristics associated with seropositivity. All estimates are corrected for test sensitivity and specificity (see Statistical appendix for details).

**Table 3.**
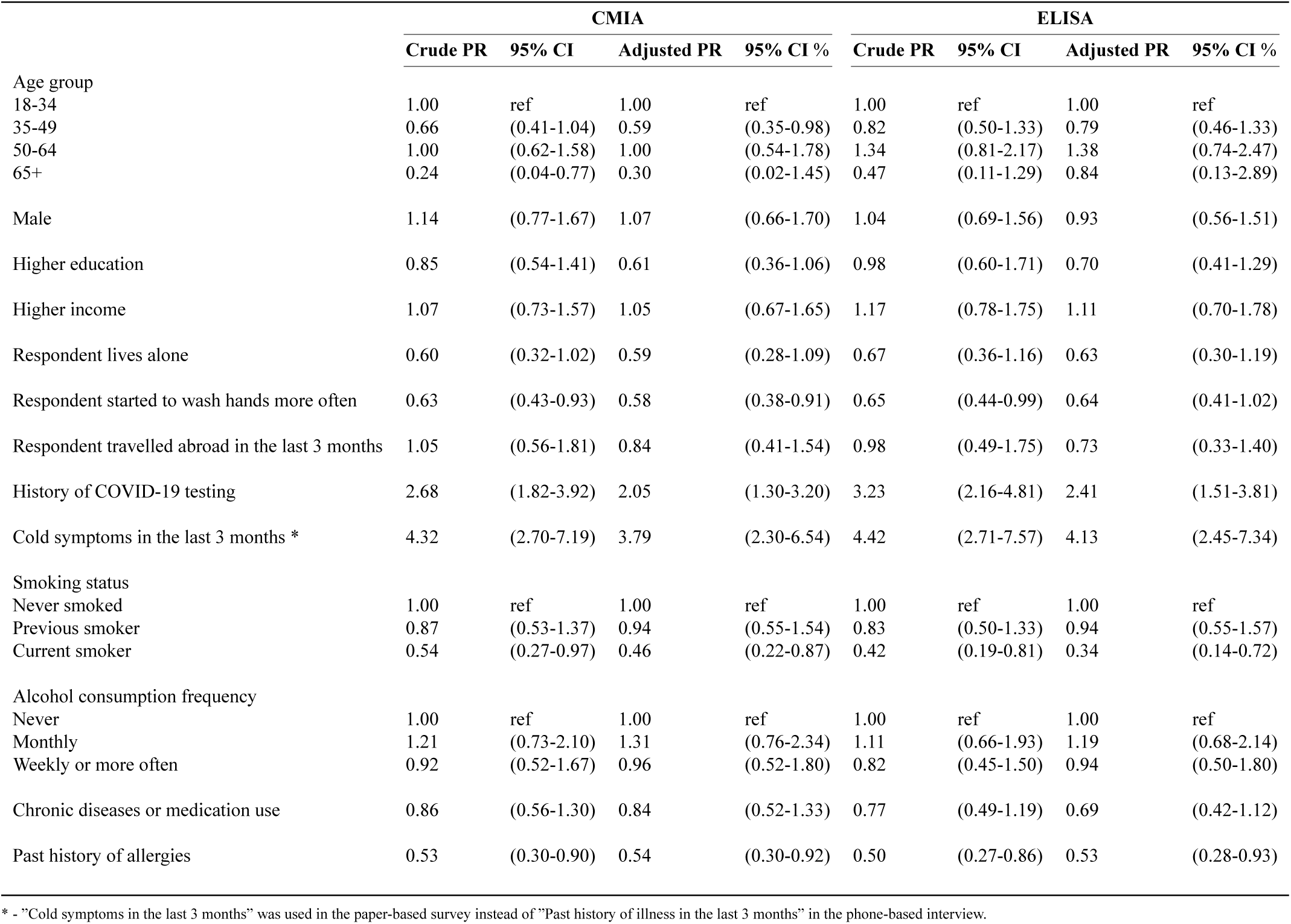
Prevalence ratios for self-reported characteristics of tested individuals in phone and paper-based surveys

|  | CMIA |  |  |  | ELISA |  |  |  |
| --- | --- | --- | --- | --- | --- | --- | --- | --- |
|  | Crude PR | 95% CI | Adjusted PR | 95% CI % | Crude PR | 95% CI | Adjusted PR | 95% CI % |
| Age group |  |  |  |  |  |  |  |  |
| 18-34 | 1.00 | ref | 1.00 | ref | 1.00 | ref | 1.00 | ref |
| 35-49 | 0.66 | (0.41-1.04) | 0.59 | (0.35-0.98) | 0.82 | (0.50-1.33) | 0.79 | (0.46-1.33) |
| 50-64 | 1.00 | (0.62-1.58) | 1.00 | (0.54-1.78) | 1.34 | (0.81-2.17) | 1.38 | (0.74-2.47) |
| 65+ | 0.24 | (0.04-0.77) | 0.30 | (0.02-1.45) | 0.47 | (0.11-1.29) | 0.84 | (0.13-2.89) |
| Male | 1.14 | (0.77-1.67) | 1.07 | (0.66-1.70) | 1.04 | (0.69-1.56) | 0.93 | (0.56-1.51) |
| Higher education | 0.85 | (0.54-1.41) | 0.61 | (0.36-1.06) | 0.98 | (0.60-1.71) | 0.70 | (0.41-1.29) |
| Higher income | 1.07 | (0.73-1.57) | 1.05 | (0.67-1.65) | 1.17 | (0.78-1.75) | 1.11 | (0.70-1.78) |
| Respondent lives alone | 0.60 | (0.32-1.02) | 0.59 | (0.28-1.09) | 0.67 | (0.36-1.16) | 0.63 | (0.30-1.19) |
| Respondent started to wash hands more often | 0.63 | (0.43-0.93) | 0.58 | (0.38-0.91) | 0.65 | (0.44-0.99) | 0.64 | (0.41-1.02) |
| Respondent travelled abroad in the last 3 months | 1.05 | (0.56-1.81) | 0.84 | (0.41-1.54) | 0.98 | (0.49-1.75) | 0.73 | (0.33-1.40) |
| History of COVID-19 testing | 2.68 | (1.82-3.92) | 2.05 | (1.30-3.20) | 3.23 | (2.16-4.81) | 2.41 | (1.51-3.81) |
| Cold symptoms in the last 3 months * | 4.32 | (2.70-7.19) | 3.79 | (2.30-6.54) | 4.42 | (2.71-7.57) | 4.13 | (2.45-7.34) |
| Smoking status |  |  |  |  |  |  |  |  |
| Never smoked | 1.00 | ref | 1.00 | ref | 1.00 | ref | 1.00 | ref |
| Previous smoker | 0.87 | (0.53-1.37) | 0.94 | (0.55-1.54) | 0.83 | (0.50-1.33) | 0.94 | (0.55-1.57) |
| Current smoker | 0.54 | (0.27-0.97) | 0.46 | (0.22-0.87) | 0.42 | (0.19-0.81) | 0.34 | (0.14-0.72) |
| Alcohol consumption frequency |  |  |  |  |  |  |  |  |
| Never | 1.00 | ref | 1.00 | ref | 1.00 | ref | 1.00 | ref |
| Monthly | 1.21 | (0.73-2.10) | 1.31 | (0.76-2.34) | 1.11 | (0.66-1.93) | 1.19 | (0.68-2.14) |
| Weekly or more often | 0.92 | (0.52-1.67) | 0.96 | (0.52-1.80) | 0.82 | (0.45-1.50) | 0.94 | (0.50-1.80) |
| Chronic diseases or medication use | 0.86 | (0.56-1.30) | 0.84 | (0.52-1.33) | 0.77 | (0.49-1.19) | 0.69 | (0.42-1.12) |
| Past history of allergies | 0.53 | (0.30-0.90) | 0.54 | (0.30-0.92) | 0.50 | (0.27-0.86) | 0.53 | (0.28-0.93) |

\* - "Cold symptoms in the last 3 months" was used in the paper-based survey instead of "Past history of illness in the last 3 months" in the phone-based interview.

**Figure 2.**
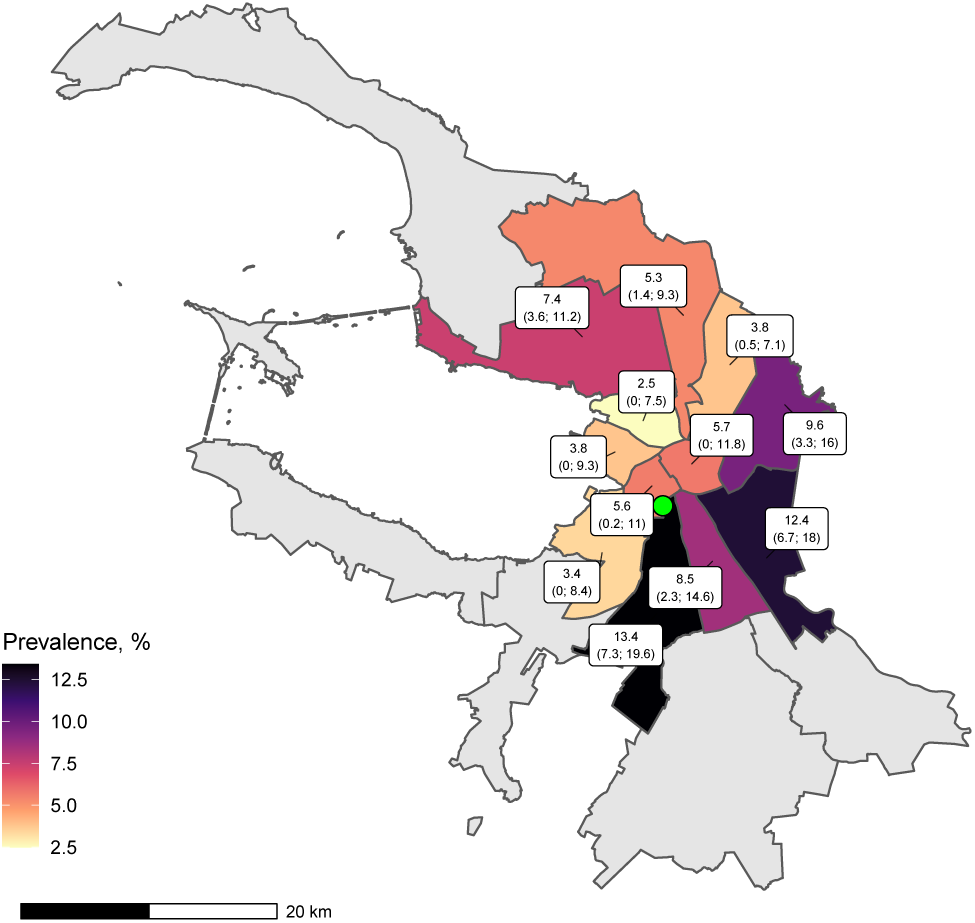
Prevalence estimates by district. This map shows CMIA-based prevalence estimates corrected for non-response bias by surveyed districts with 95% CIs in parentheses. Green dot is the clinic test site location. Remote districts excluded from survey are in grey. Map data copyrighted OpenStreetMap contributors.

We observed a slight increase in seroprevalence by the week of the phone interview (see Figure 3a) and by the week of the blood draw (see Figure 3b).

**Figure 3.**
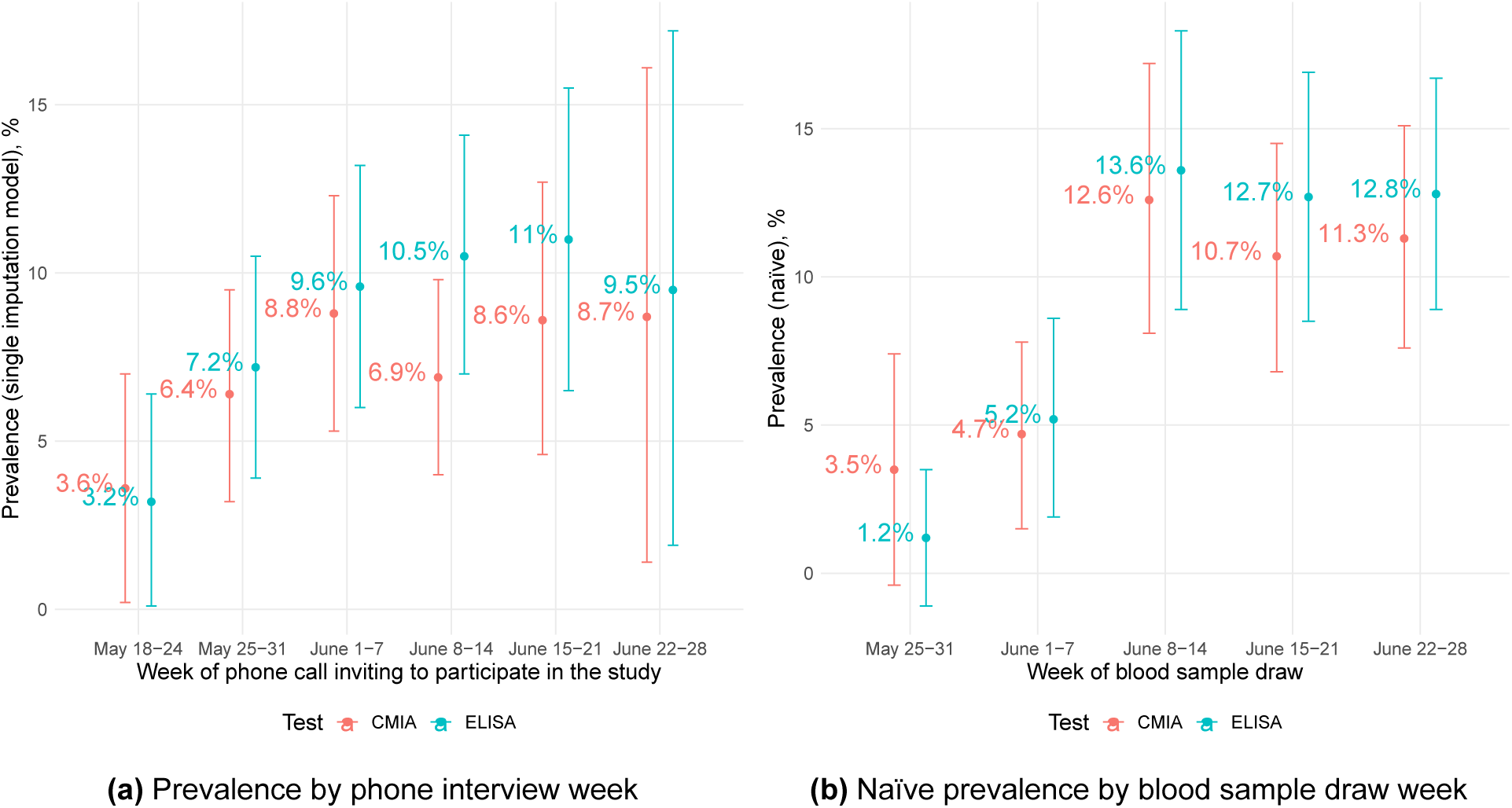
Prevalence estimates over time.

Our secondary analysis of participants who filled out clinic paper-based survey forms revealed additional covariates associated with seroconversion. It was negatively associated with smoking status with prevalence ratios 0.46 (95%CI 0.22-0.87) and 0.34 (95%CI 0.14-0.72) (PR for current smokers vs non-smokers based on CMIA and ELISA, respectively), and self-reported history of allergies with prevalence ratios 0.54 (95%CI 0.30-0.90) and 0.53 (95%CI 0.28-0.93). (see Supplementary Appendix Table 3).

### Sensitivity analysis

Alternative definitions of seroprevalence (test combination either favouring sensitivity or specificity) did not qualitatively change the effect of non-response bias (see Supplementary Appendix Table A4). Seroprevalence estimates obtained on re-weighted survey data (based on age group and education attainment level) were similar to estimates from the main analysis. (see Supplementary Appendix Table A5).

## Discussion

Our study aimed to assess the spread of epidemic in the fourth largest European city — St. Petersburg. Although the seroprevalence estimate varied based on the test used and type of correction applied, the total number of population with detectable antibodies was still far lower than the proportion needed for herd immunity. Overall seroprevalence in the range between 7% and 10% was in line with the results obtained from the previous studies and provides evidence of the similar epidemic development across the world with less than one tenth of population affected in the first months [5; 6].

To the best of our knowledge, this is the first seroprevalence survey of COVID-19 that applied correction based on characteristics that are associated with the risk of seropositivity in combination with incentivised participation. Early COVID-19 serological surveys are likely to exhibit high sampling error because of recruitment methods.[22] Population based studies with random sampling relied on probability weighting obtained from the comparison with the source population [5–7].. Our findings show that even low estimates of seroprevalence (around or below 10%) obtained in population surveys can be an overestimation in populations with high risk of non-response bias.

We detected only a slight change in the estimate of seroprevalence when we corrected our estimated for non-response bias with respect to demographic or socioeconomic characteristics, but far more significant difference was detected when several behavioural characteristics were included in models and applied in the correction. In general, our analysis shows that naïve estimates that do not account for the non-response bias tend to drive prevalence estimates upward. In contrast to the findings in the literature examining the non-response bias in HIV serosurveys, on average participants who are more likely to have antibodies are more likely to participate in COVID-19 surveys [16; 23]. Participants with history of illness in the last 3 months or past history of tests for COVID-19 in the last 3 months were more likely to agree to antibody testing in our study probably seeking external confirmation.

In our sample of participants we did find only a slight age difference in the seropositivity rates, and there was no difference between men and women, which is in line with previous findings [6]. However, we observed several clear differences in seroprevalence estimates in a subgroup analysis. First of all, we detected an elevated seroprevalence in participants who reported history of illness and history of any COVID-19 test in the last 3 months, this association was seen regardless of the modelling approach. Second, seroprevalence was lower in participants who lived alone and reported that they started to wash their hands more often. Third, in the secondary analysis of participants who were tested we observed that seroprevalence was lower in current smokers compared to never smokers, it was also lower in participants who reported past history of allergies.

All associations revealed in our study should not be immediately regarded as causal due to limitations in the study design and analysis. History of testing and illness in the last 3 months can be easily interpreted. Seroprevalence among those reporting a history of COVID-19 testing was relatively low (around 20%), this can be explained by the high scale of testing in Russia since the onset of the epidemic. However, our study is not a direct evidence of the effectiveness of hand hygiene, as self-reported change in habits can reflect other differences between sub-populations. There is limited and conflicting evidence about the smoking rates in COVID-19 patients [24; 25]. While our study is the one of the first that compared population-based seroprevalence estimates between smokers and non-smokers there is a need for more studies to confirm this finding [9]. There are many examples when smoking effects were subject to structural epidemiological biases [26]. Even if this association is causal, then behavioural or biological mechanisms should be explored. Smoking is a well-established risk factor for many diseases and it is likely linked to COVID-19 severity regardless of the risk of infection [24].

It is also tempting to immediately search for biological explanation that link allergy status and risk of infection [27]. However, we should be very cautious due to limitations of study design and other possible explanations, e.g. people who self-report being allergic may behave in a way to minimize risk of being infected. The question about allergy was very general in our paper-based survey, that also limits the value of this finding.

Important source of bias in serological studies is the performance and the nature of the serological tests [28]. Possible explanation of the difference in our study includes different classes of Ig analysed — IgG in case of CMIA and IgG+IgM+IgA in case of ELISA. However, given the total seroprevalence of not more than 10% it seems that lack of IgM and IgA in CMIA test can only partially explain the difference. A recent study showed that seroconversion started on day 5 after disease onset and IgG level rose even earlier than IgM [29]. Another possible explanation for different seroprevalence estimates of two tests is the nature of antigen. SARS-CoV-2 antibody responses specific to the Spike (S) and/or the nucleocapsid (N) proteins are equally sensitive in the acute infection phase [30]. However, as compared to anti-S antibody responses, those against the N protein appear to wane in the post-infection [31]. Recent evaluations of CMIA test used in our study reported sensitivity far below 100% reported by manufacturer. This may also explain the difference [32; 33]. Another source of underestimation is a proportion of infected that do not seroconvert. Straightforward adjustments for this sort of biases are not available without additional laborious testing [34].

Our study has several other important limitations. We are addressing seroprevalence in adults only, while previous studies also included participants younger than 18 years old [5; 6]. Our study had a relatively low participation rate given the existing propensity to answer phone calls in the city. However, we assumed missingness at random for those who did not complete the interview or did not pick the phone. Comparison with the previous representative city survey showed that our sample was representative (see Supplementary Appendix Table A3). We have also excluded distant city districts from our sampling. Even though we observed statistically significant differences between by-district seroprevalence, the lion’s share of city residents (about 4.3 mln of 5.2 mln) live in the surveyed districts. Our randomized incentivisation scheme was not successful because randomly assigned taxi offer was not associated with participation agreement and failed to become a valid exclusion restriction. In our main analysis we did not apply post-stratification methods adopted previously [5]. However, application of raking weights estimated to match targets from a representative survey of adult city population showed little to no changes in weighted seroprevalence estimates. We explained this by little to no association between seroconversion and age or education level. Finally, we report cross-sectional results but longitudinal data are needed to offer additional insights to immunity waning and prolonged defence against re-infection.

### Conclusion

COVID-19 pandemic has already affected at least 300 000 residents of St. Petersburg that can be extrapolated to millions in the whole country. However the vast majority of population does not carry antibodies to SARS-CoV-2. This highlights the need for further high-quality population based studies that can provide evidence for measures to diminish the impact of the pandemic.

## Data Availability

Study data and code is available online (https:// github.com/eusporg/spb_covid_study20).

## Authors’ contributions

AB, DSk, VV, KT, LB, DSh and PT conceived the study. AB, DSk and VV drafted the first version of the manuscript. KT, YR, AN, EP and DSh contributed to drafting sections of the manuscript. DS, AB and DSh did data analyses. SZ and EP did lab analyses. All authors participated in the study design, helped to draft the manuscript, contributed to the interpretation of data and read and approved the final manuscript.

## Acknowledgements

We acknowledge personal support from Vitaly Nesis (Chief Executive Officer, Polymetal International, plc). We thank Alla Samoletova (European University at St. Petersburg) for administrative support and management of the study. We are also beholden to Dmitriy Serebrennikov (EU SPb) for managing paper-based survey data entry, Ruslan Kuchakov (EU SPb) for initial assistance with visualizations. We also gratefully acknowledge support from Yana Novikova and Aleksey Gladkikh (Invitro Laboratory) regarding the CMIA testing, Yulia Stepantsova (Chursina) regarding phone based interviewers, Maya Perestoronina (Clinic “Scandinavia”) for comments on the protocol, Lizaveta Dubovik and Irina Shubina for the science communication, and Sergey Nechiporenko for the protocol translation. We thank the interviewers, nurses, general practitioners, and administrative personnel of the Clinic “Scandinavia”. We also thank Ilya Fomintsev for his help and support during the initial stages of the study. We also thank all study participants.

## Supplementary materials

### Statistical appendix

Let adult population of St. Petersburg be of size *N*, indexed with *i*, and characterised by a triplet ⟨ *Z, D, Y* ⟩. *Z*_*i*_ = 1 means that the *i*-th resident will be surveyed, with *Z*_*i*_ = 0 otherwise. Let **Z** be a vector of size *N*, its *i*-th element equals *Z*_*i*_. In other words, **Z** indexes which city residents will be surveyed.

*D* marks the individual decision to participate in the survey when contacted: *D*_*i*_(**Z**) = 1: *i*-th individual volunteers to take part in the study, with *D*_*i*_(**Z**) = 0 otherwise. Let **D** be the volunteer vector of size *N*.

When participation is universal *D*_*i*_(**Z**) = *Z*_*i*_ ∀*i*. In reality one could observe non-zero refusal rates. In what follows we assume one-sided noncompliance.

Variable *Y* characterises antibody status (seroconversion) of the *i*-th individual and takes the following values: { 1, 0 }, where 1 — has antibodies to SARS-CoV-2 and 0 — does not have antibodies. Antibody status can be both observed and unobserved: *Y*_*i*_ ≡ *Y*_*i*_ (**Z, D**). We are able to observe antibody status for *Y*_*i*_ (*Z*_*i*_ = 1, *D*_*i*_ = 1), i.e. for the surveyed individuals who agreed to volunteer in the study and were tested.

We are interested in population seroprevalence estimate 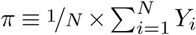 Having conducted our study we can estimate the naïve seroprevalence 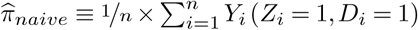, where *n* is the number of tested individuals. To arrive at population-level seroprevalence estimates the following assumptions are required:

### Random sampling

The survey is conducted such that Pr (**Z** = **c**) = Pr **Z** = **c**′ where **c** and **c**′ are arbitrary survey vectors.

### Stable unit treatment value assumption (SUTVA)

Both observed and unobserved individual antibody status depends only on his/her antibody status and decision to participate in the survey and not on the decision of other study participants. The surveyed individuals maintain no contacts or other social interactions that could influence their individual decisions to participate in the study. Such interactions might arise in case of household sampling where all household members are invited to participate in the survey.

### No unobservable characteristics of study participants

Let **X** ∈ℝ^*N×M*^ be a matrix of observable individual characteristics. Its row **X**_*i*_ stores all characteristics of individual *i* that influence his/her decision to participate in the study when surveyed. Naïve seroprevalence estimate will be representative of the city population only when cov ([D | X], [Z| X]) = 0 and cov ([D| X], [Y| X]) = 0. That is, conditional on all observable characteristics of individuals, the decision to participate in the study is orthogonal to both the conditional-on-observables survey inclusion and conditional-on-observables antibody status. While the former orthogonality assumption [*D*| *X*] ⊥ [*Z*| *X*] is satisfied under random sampling of individuals, the latter is more problematic. First, no phone survey design could ensure full observability of all characteristics associated both with individual decision to volunteer and antibody status. Many variables might be left unobserved, rendering this orthogonality assumption unrealistic. Second, differing sets of observables can influence seroconversion and decision to volunteer. For instance, past travel history may be an important observable responsible for *D* = 1 while fully independent of actual antibody status *Y* = 1.

When the above assumptions are not satisfied the naïve seroprevalence estimate is biased. To account for this we rely on bivariate binary model with non-random selection in the spirit of Bärnighausen et al. (2011) [1]. First we assume that decision of a surveyed individual to agree to participate in the survey and come to the clinic test site is determined by a latent variable

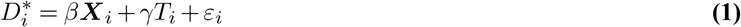

such that *D*_*i*_ = 1 if *D*^*∗*^*>* 0 and *D*_*i*_ = 0 otherwise. *T*_*i*_ is a variable equal to unity if surveyed individual was offered free taxi to and from the clinic test site during the phone survey, *ε*_*i*_ is the error term. We observe *D* only for *n* out of *N* individuals in the population.

We observe antibody status only for those with *D*_*i*_ = 1 and assume that seroconversion is determined by a latent variable

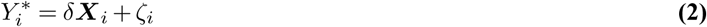

such that *Y*_*i*_ = 1 if *Y* ^*∗*^*>* 0 and *Y*_*i*_ = 0 otherwise. *ζ*_*i*_ is the error term. Since we have offered taxi to random phone survey participants we can safely assume that cov ([Y|X], T) = 0 and *T* becomes a valid exclusion restriction.

We impose a structural assumption of independent and identically Normal-distributed error terms *ε*_*i*_ and *ζ*_*i*_ with nil mean and unit variance. Their joint cumulative distribution function is given by Φ(*ε*_*i*_, *ζ*_*i*_, *ρ*) where *ρ* is covariance (correlation coefficient).

This bivariate probit is estimated with R package GJRM [2]. We use different definitions of seropositivity *Y* = 1 depending on antibody tests or combinations thereof and different set of variables in design matrix **X**.

Our first step is to estimate 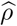 and test whether it is statistically significantly different from zero. Estimates under different sets of variables are reported below with 95% CIs in parentheses are reported in Supplementary Appendix Table A1.

As our baseline we adopt a model where a rich set of demographic, socioeconomic, and seropositivity-related characteristics is included in the design matrix **X**. In this model results from both the simulated CIs and Lagrange multiplier test with null *ρ* = 0 (not reported here, available at request) suggest that one cannot reject the null hypothesis of error term independence between the selection stage and antibody test result stage.

Under error term independence Heckman correction is not required to arrive at seroprevalence estimates for the entire city population when response is non-random. However, the naïve seroprevalence estimate can still be biased since the tested individuals are not representative of the city population. To circumvent this we use the estimated parameters from baseline seroconversion probit (see equation 2 above) and predict antibody status *Y* for all surveyed individuals regardless of their agreement to participate in the survey. Such (univariate) single imputation that assumes no unobserved confounders permits us to correct the naïve seroprevalence estimates for missing data for those individuals who have refused to get tested or did not visit the clinic.

Symmetric confidence intervals come from standard errors estimated with delta method. Results do not change qualitatively when we consider non-symmetric confidence intervals after Bayesian posterior simulation of the parameter vector estimate (not reported here, available at request).

Finally, we rely on manufacturers’ test characteristics to correct all reported prevalence estimates 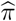 using the formula

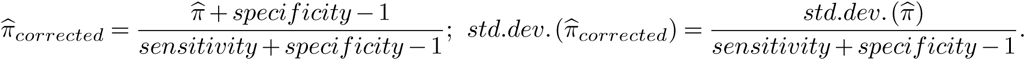

**Table A1.** Estimated error term correlation between selection stage and testing stage

| Design matrix **X** | $\hat{\rho}$, CMIA | $\hat{\rho}$, ELISA |
| --- | --- | --- |
| Demographic characteristics | -0.71 (-0.74;-0.68) | -0.71 (-0.74;-0.67) |
| Demographic and socioeconomic characteristics | -0.69 (-0.73;-0.67) | -0.69 (-0.72;-0.66) |
| Characteristics associated with seropositivity | 0.32 (-1.0;1.0) | -0.94 (-1.0;-0.09) |
| Demographics, socioeconomic status and characteristics associated with seropositivity | 0.50 (-1.0;1.0) | 0.66 (-0.95;0.99) |

## STROBE checklist for cross-sectional studies

|  | Item No | Recommendation | Page |
| --- | --- | --- | --- |
| Title and abstract |  |  |  |
|  | 1a | Indicate the study's design with a commonly used term in the title or the abstract | See page 1 |
|  | 1b | Provide in the abstract an informative and balanced summary of what was done and what was found | See page 1 |
| Introduction |  |  |  |
| Background/rationale | 2 | Explain the scientific background and rationale for the investigation being reported | See page 1 |
| Objectives | 3 | State specific objectives, including any prespecified hypotheses | See page 2 |
| Methods |  |  |  |
| Study design | 4 | Present key elements of study design early in the paper | See page 2 |
| Setting | 5 | Describe the setting, locations, and relevant dates, including periods of recruitment, exposure, follow-up, and data collection | See page 2 |
| Participants | 6 | Give the eligibility criteria, and the sources and methods of selection of participants | See page 2 |
| Variables | 7 | Clearly define all outcomes, exposures, predictors, potential confounders, and effect modifiers. Give diagnostic criteria, if applicable | See page 2 |
| Data sources/ measurement | 8 | For each variable of interest, give sources of data and details of methods of assessment (measurement). Describe comparability of assessment methods if there is more than one group | See pages 2 |
| Bias | 9 | Describe any efforts to address potential sources of bias | See page 2 |
| Study size | 10 | Explain how the study size was arrived at | See page 2 |
| Quantitative variables | 11 | Explain how quantitative variables were handled in the analyses. If applicable, describe which groupings were chosen and why | See page 3 |
| Statistical methods | 12a | Describe all statistical methods, including those used to control for confounding | See page 3 |
| Statistical methods | 12b | Describe any methods used to examine subgroups and interactions | See page 3 |
| Statistical methods | 12c | Explain how missing data were addressed | See page 3 |
| Statistical methods | 12d | If applicable, describe analytical methods taking account of sampling strategy | See page 3 |
| Statistical methods | 12e | Describe any sensitivity analyses | See page 3 |
| Results |  |  |  |
| Participants | 13a | Report numbers of individuals at each stage of study – eg numbers potentially eligible, examined for eligibility, confirmed eligible, included in the study, completing follow-up, and analysed | See page 3 |
| Participants | 13b | Give reasons for non-participation at each stage | See page 3 |
| Participants | 13c | Consider use of a flow diagram | See page 3 |
| Descriptive data | 14a | Give characteristics of study participants (eg demographic, clinical, social) and information on exposures and potential confounders | See page 3 |
| Descriptive data | 14b | Indicate number of participants with missing data for each variable of interest | See page 3 |
| Outcome data | 15 | Report numbers of outcome events or summary measures | See page 4 |
| Main results | 16a | Give unadjusted estimates and, if applicable, confounder-adjusted estimates and their precision (eg, 95% confidence interval). Make clear which confounders were adjusted for and why they were included | See page 4 |
| Main results | 16b | Report category boundaries when continuous variables were categorised | NA |
| Main results | 16c | If relevant, consider translating estimates of relative risk into absolute risk for a meaningful time period | NA |
| Other analyses | 17 | Report other analyses done—eg analyses of subgroups and interactions, and sensitivity analyses | See page 4 |
| Discussion |  |  |  |
| Key results | 18 | Summarise key results with reference to study objectives | See page 4 |
| Limitations | 19 | Discuss limitations of the study, taking into account sources of potential bias or imprecision. Discuss both direction and magnitude of any potential bias | See page 5 |
| Interpretation | 20 | Give a cautious overall interpretation of results considering objectives, limitations, multiplicity of analyses, results from similar studies, and other relevant evidence | See page 4 |
| Generalisability | 21 | Discuss the generalisability (external validity) of the study results | See page 4 |
| Other information |  |  |  |
| Funding | 22 | Give the source of funding and the role of the funders for the present study and, if applicable, for the original study on which the present article is based | See page 5 |

**Figure A1.**
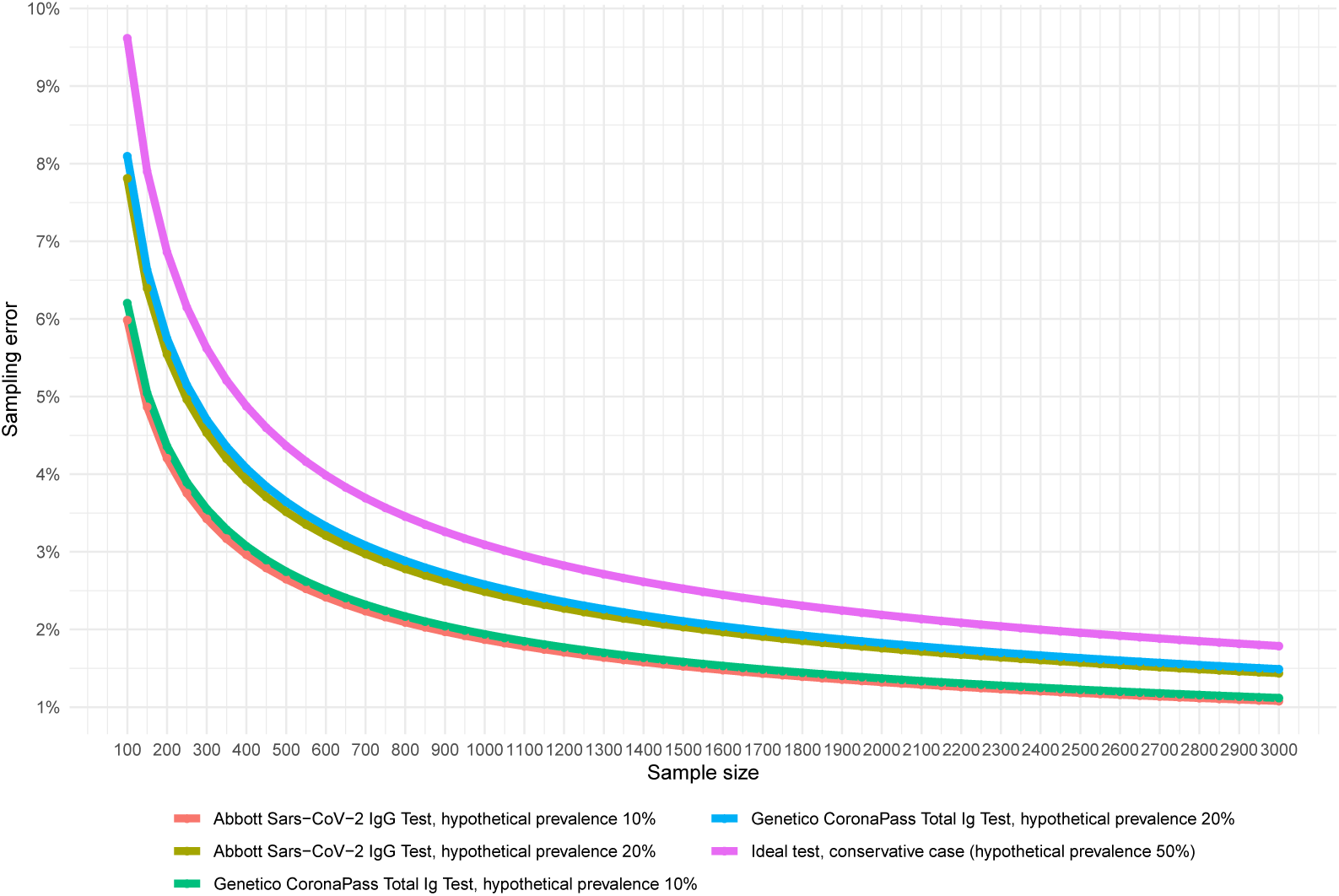
Sampling error under different test characteristics. This chart reports the relationship between sampling error and sample size under different test characteristics and assumed hypothetical prevalence in the population. Calculations are made with the designated code [3].

**Table A2.** Summary statistics of eligible phone survey respondents

|  |  | Overall | Not tested | Tested |
| --- | --- | --- | --- | --- |
| Number of participants |  | 6400 | 5362 | 1038 |
| Age group |  |  |  |  |
|  | 18–34 | 2333 (36.5) | 1937 (36.1) | 396 (38.2) |
|  | 35–49 | 2036 (31.8) | 1679 (31.3) | 357 (34.4) |
|  | 50–64 | 1271 (19.9) | 1053 (19.6) | 218 (21.0) |
|  | 65+ | 760 (11.9) | 693 (12.9) | 67 (6.5) |
| Sex |  |  |  |  |
|  | Female | 3766 (58.8) | 3114 (58.1) | 652 (62.8) |
|  | Male | 2634 (41.2) | 2248 (41.9) | 386 (37.2) |
| Higher education |  |  |  |  |
|  | No | 2108 (32.9) | 1929 (36.0) | 179 (17.2) |
|  | Yes | 4292 (67.1) | 3433 (64.0) | 859 (82.8) |
| Higher income |  |  |  |  |
|  | No | 3484 (54.4) | 2988 (55.7) | 496 (47.8) |
|  | Yes | 2579 (40.3) | 2076 (38.7) | 503 (48.5) |
|  | NA | 337 (5.3) | 298 (5.6) | 39 (3.8) |
| Respondent lives alone |  |  |  |  |
|  | No | 5206 (81.3) | 4363 (81.4) | 843 (81.2) |
|  | Yes | 1194 (18.7) | 999 (18.6) | 195 (18.8) |
| Respondent travelled abroad in the past 3 months |  |  |  |  |
|  | No | 5982 (93.5) | 5064 (94.4) | 918 (88.4) |
|  | Yes | 418 (6.5) | 298 (5.6) | 120 (11.6) |
| Respondent started to wash hands more often |  |  |  |  |
|  | No | 2190 (34.2) | 1893 (35.3) | 297 (28.6) |
|  | Yes | 4155 (64.9) | 3419 (63.8) | 736 (70.9) |
|  | NA | 55 (0.9) | 50 (0.9) | 5 (0.5) |
| History of illnesses in the past 3 months |  |  |  |  |
|  | No | 4324 (67.6) | 3747 (69.9) | 577 (55.6) |
|  | Yes | 1997 (31.2) | 1543 (28.8) | 454 (43.7) |
|  | NA | 79 (1.2) | 72 (1.3) | 7 (0.7) |
| History of COVID 19 testing |  |  |  |  |
|  | No | 5425 (84.8) | 4621 (86.2) | 804 (77.5) |
|  | Yes | 975 (15.2) | 741 (13.8) | 234 (22.5) |
| City district |  |  |  |  |
|  | Admiralteyskiy | 265 (4.1) | 205 (3.8) | 60 (5.8) |
|  | Frunzenskiy | 477 (7.5) | 403 (7.5) | 74 (7.1) |
|  | Kalininskiy | 734 (11.5) | 616 (11.5) | 118 (11.4) |
|  | Kirovskiy | 437 (6.8) | 384 (7.2) | 53 (5.1) |
|  | Krasnogvardeyskiy | 534 (8.3) | 457 (8.5) | 77 (7.4) |
|  | Moskovskiy | 538 (8.4) | 427 (8.0) | 111 (10.7) |
|  | Nevskiy | 842 (13.2) | 717 (13.4) | 125 (12.0) |
|  | Petrogradskiy | 192 (3.0) | 157 (2.9) | 35 (3.4) |
|  | Primorskiy | 955 (14.9) | 789 (14.7) | 166 (16.0) |
|  | Tsentralniy | 342 (5.3) | 290 (5.4) | 52 (5.0) |
|  | Vasileostrovskiy | 250 (3.9) | 207 (3.9) | 43 (4.1) |
|  | Vyborgskiy | 834 (13.0) | 710 (13.2) | 124 (11.9) |
| Week of phone interview |  |  |  |  |
|  | May 18-24 | 473 (7.4) | 372 (6.9) | 101 (9.7) |
|  | May 25-31 | 1333 (20.8) | 1110 (20.7) | 223 (21.5) |
|  | June 1-7 | 1419 (22.2) | 1190 (22.2) | 229 (22.1) |
|  | June 8-14 | 1496 (23.4) | 1231 (23.0) | 265 (25.5) |
|  | June 15-21 | 981 (15.3) | 817 (15.2) | 164 (15.8) |
|  | June 22-28 | 698 (10.9) | 642 (12.0) | 56 (5.4) |
| CMIA positive test |  |  |  |  |
|  | No | 941 (14.7) | 0 (0.0) | 941 (90.7) |
|  | Yes | 97 (1.5) | 0 (0.0) | 97 (9.3) |
|  | NA | 5362 (83.8) | 5362 (100.0) | 0 (0.0) |
| ELISA positive test |  |  |  |  |
|  | No | 928 (14.5) | 0 (0.0) | 928 (89.4) |
|  | Yes | 107 (1.7) | 0 (0.0) | 107 (10.3) |
|  | NA | 5365 (83.8) | 5362 (100.0) | 3 (0.3) |
| Smoking status (*paper survey*) |  |  |  |  |
|  | Never smoked | 462 (7.2) | 0 (0.0) | 462 (44.5) |
|  | Used to smoke | 263 (4.1) | 0 (0.0) | 263 (25.3) |
|  | Smoking now | 195 (3.0) | 0 (0.0) | 195 (18.8) |
|  | NA | 5480 (85.6) | 5362 (100.0) | 118 (11.4) |
| Past history of allergies (*paper survey*) |  |  |  |  |
|  | No | 631 (9.9) | 0 (0.0) | 631 (60.8) |
|  | Yes | 277 (4.3) | 0 (0.0) | 277 (26.7) |
|  | NA | 5492 (85.8) | 5362 (100.0) | 130 (12.5) |
| Chronic diseases or medication use (*paper survey*) |  |  |  |  |
|  | No | 534 (8.3) | 0 (0.0) | 534 (51.4) |
|  | Yes | 418 (6.5) | 0 (0.0) | 418 (40.3) |
|  | NA | 5448 (85.1) | 5362 (100.0) | 86 (8.3) |
| Cold symptoms in the past 3 months (*paper survey*) |  |  |  |  |
|  | No | 532 (8.3) | 0 (0.0) | 532 (51.3) |
|  | Yes | 418 (6.5) | 0 (0.0) | 418 (40.3) |
|  | NA | 5450 (85.2) | 5362 (100.0) | 88 (8.5) |
| Alcohol consumption frequency (*paper survey*) |  |  |  |  |
|  | Never | 218 (3.4) | 0 (0.0) | 218 (21.0) |
|  | Monthly | 404 (6.3) | 0 (0.0) | 404 (38.9) |
|  | Weekly or more often | 318 (5.0) | 0 (0.0) | 318 (30.6) |
|  | NA | 5460 (85.3) | 5362 (100.0) | 98 (9.4) |

**Table A3.** Sample means across study stages and in relation to a representative survey

|  | Called | Agreed | Tested | KOUZh-2018 |
| --- | --- | --- | --- | --- |
| N | 6384 | 3370 | 1028 | 2977 |
| N (paper-based survey) |  |  | 911 |  |
| Male, % | 41.3 (40.1-42.5) | 37.4 (35.7-39) | 37.1 (34.1-40) | 40.2 (38.4-41.9) |
| Age, years | 43.1 (42.7-43.5) | 42.1 (41.6-42.6) | 41.4 (40.6-42.3) | 46.7 (46.1-47.3) |
| 18-34, % | 36.3 (35.1-37.4) | 37.7 (36.1-39.4) | 38.2 (35.3-41.2) | 27.9 (26.3-29.5) |
| 35-49, % | 31.8 (30.7-33.0) | 33 (31.4-34.6) | 34 (31.1-36.9) | 31.7 (30-33.3) |
| 50-64, % | 20.0 (19.0-20.9) | 19.9 (18.6-21.3) | 21.2 (18.7-23.7) | 22.8 (21.3-24.3) |
| 65+, % | 12 (11.2-12.8) | 9.3 (8.4-10.3) | 6.5 (5.0-8.0) | 17.6 (16.3-19.0) |
| Education |  |  |  |  |
| Primary education, % | 1.1 (0.8-1.4) | 0.8 (0.0-0.6) | 0.3 (0.5-1.1) | 2.7 (2.1-3.3) |
| Complete secondary education, % | 8.9 (8.2-9.6) | 7.5 (6.6-8.4) | 5.0 (3.6-6.3) | 10.2 (9.1-11.3) |
| Special secondary education, % | 23.2 (22.1-24.2) | 20.1 (18.8-21.5) | 11.9 (9.9-13.8) | 39.5 (37.7-41.3) |
| Higher education, % | 66.9 (65.7-68.0) | 71.6 (70.1-73.1) | 82.9 (80.6-85.2) | 47.6 (45.8-49.4) |
| Employed, % | 68.3 (67.1-69.4) | 71.1 (69.6-72.7) | 78.7 (76.2-81.2) | 70.2 (68.6-71.8) |
| Self-reported health status |  |  |  |  |
| Very good, % | 19.9 (18.9-20.8) | 16.9 (15.6-18.1) | 15.2 (13-17.4) | 7.8 (6.8-8.7) |
| Good, % | 48.9 (47.6-50.1) | 50.6 (48.9-52.3) | 53.5 (50.4-56.6) | 45.3 (43.5-47.1) |
| Satisfactory, % | 28.5 (27.4-29.6) | 29.8 (28.2-31.3) | 29.7 (26.9-32.5) | 39.2 (37.5-41) |
| Bad, % | 2.4 (2.1-2.8) | 2.4 (1.9-3.0) | 1.6 (0.8-2.3) | 7.1 (6.2-8.0) |
| Very bad, % | 0.3 (0.2-0.5) | 0.3 (0.1-0.5) | 0.1 (0.0-0.3) | 0.6 (0.3-0.9) |
| Lives alone, % | 18.8 (17.8-19.7) | 18.1 (16.8-19.4) | 19 (16.6-21.4) | 19.7 (18.3-21.1) |
| Has cellphone, % | 100.0 (100.0-100.0) | 100.0 (100.0-100.0) | 100 (100.0-100.0) | 99.5 (99.3-99.8) |
| Self-reported smoking status (from paper-based survey) |  |  |  |  |
| Never smoked, % |  |  | 49.9 (46.7-53.2) | 52.9 (51.1-54.7) |
| Smoking earlier, % |  |  | 28.9 (25.9-31.8) | 15.5 (14.2-16.8) |
| Smoking now, % |  |  | 21.2 (18.5-23.8) | 31.6 (30.0-33.3) |
| Consumes alcohol, % |  |  | 76.6 (73.9-79.4) | 74. (73.1-76.2) |

95% confidence intervals in parentheses. "Called" means individuals who agreed to participate in the phone survey-"Agreed" are individuals who agreed to be contacted by the clinic to get tested-"Tested" are individuals who came to clinic and gave blood samples. KOUZh-2018 is the 2016 round of the Comprehensive monitoring of living conditions household survey carried out by the Federal State Statistics Service of Russia. We subset this survey to include only adults in St. Petersburg. We report only complete-case observations in terms of all variables, therefore the number of observations is slightly lower due to listwise deletion.

**Table A4.** SARS-CoV-2 seroprevalence estimates from bivariate probit models with different sets of individual characteristics for non-response bias correction and alternative definitions of seropositivity

| Regressors included in bivariate probit model | CMIA or ELISA |  |  |  | CMIA and ELISA |  |  |  |
| --- | --- | --- | --- | --- | --- | --- | --- | --- |
|  | Number of participants |  | Seroprevalence (95% CI) |  | Number of participants |  | Seroprevalence (95% CI) |  |
|  | Interviewed | Tested | Naïve | Single imputation | Interviewed | Tested | Naïve | Single imputation |
| Demographic characteristics | 6400 | 1038 | 11.2% (9.3-13.2) | 10.9% (9.0-12.8) | 6397 | 1035 | 8.2% (6.5-9.9) | 8.0% (6.3-9.6) |
| Demographic and socioeconomic characteristics | 6063 | 999 | 11.6% (9.6-13.6) | 11.5% (9.3-13.6) | 6061 | 997 | 8.5% (6.7-10.2) | 8.3% (6.4-10.2) |
| Characteristics associated with seropositivity | 6267 | 1026 | 11.3% (9.3-13.2) | 9.2% (7.5-10.9) | 6264 | 1023 | 8.2% (6.5-10.0) | 6.6% (5.0-8.1) |
| Demographics, socioeconomic status and characteristics associated with seropositivity | 5953 | 990 | 11.6% (9.6-13.6) | 9.7% (7.7-11.6) | 5951 | 988 | 8.4% (6.7-10.2) | 6.8% (5.2-8.5) |

"Demographic characteristics" means the following variables: individual age group (18-34, 35-49, 50-64, 65+ years old) and sex. "Socioeconomic characteristics" means the following variables: higher education status and higher self-reported income level. "Characteristics associated with seropositivity" means the following variables: history of illness in the last 3 months, history of COVID-19 testing, whether respondent lives alone, change in hand washing habits during pandemic, week of the phone interview, and city district. All models include a variable indicating random offer of taxi transportation to and from the clinic test site for interviewed participants. All estimates are corrected for tests characteristics.

**Table A5.** SARS-CoV-2 seroprevalence estimates from bivariate probit models after re-weighting the sample with raking weights (age group and educational attainment level) estimated to match a 2016 representative survey of adult city residents

| Regressors included in bivariate probit model | CMIA |  |  |  | ELISA |  |  |  |
| --- | --- | --- | --- | --- | --- | --- | --- | --- |
|  | Number of participants |  | Seroprevalence (95% CI) |  | Number of participants |  | Seroprevalence (95% CI) |  |
|  | Interviewed | Tested | Naïve | Single imputation | Interviewed | Tested | Naïve | Single imputation |
| Demographic characteristics | 6400 | 1038 | 9.0% (7.2-10.8) | 8.5% (6.7-10.4) | 6397 | 1035 | 10.5% (8.6-12.3) | 9.6% (7.8-11.4) |
| Demographic and socioeconomic characteristics | 6063 | 999 | 9.3% (7.5-11.1) | 8.9% (6.4-11.3) | 6061 | 997 | 10.8% (8.9-12.8) | 10.6% (7.9-13.2) |
| Characteristics associated with seropositivity | 6267 | 1026 | 9.0% (7.2-10.8) | 6.7% (5.2-8.2) | 6264 | 1023 | 10.5% (8.6-12.4) | 8.2% (6.6-9.9) |
| Demographics, socioeconomic status and characteristics associated with seropositivity | 5953 | 990 | 9.3% (7.5-11.2) | 7.2% (5.1-9.3) | 5951 | 988 | 10.8% (8.9-12.8) | 8.8% (6.5-11.0) |

“Demographic characteristics” means the following variables: individual age group (18-34, 35-49, 50-64, 65+ years old) and sex. “Socioeconomic characteristics” means the following variables: higher education status and higher self-reported income level. “Characteristics associated with seropositivity” means the following variables: history of illness in the last 3 months, history of COVID-19 testing, whether respondent lives alone, change in hand washing habits during pandemic, week of the phone interview, and city district. All models include a variable indicating random offer of taxi transportation to and from the clinic test site for interviewed participants. All estimates are corrected for tests characteristics. Serosurvey sample was re-weighted with raking weights estimated to match the survey age group and educational attainment proportions in 2016 representative survey of adult city population (see Supplementary Appendix Table A3 for description of this survey and the target proportions). R package anesrake was used to compute the weights.

**Figure A2.**
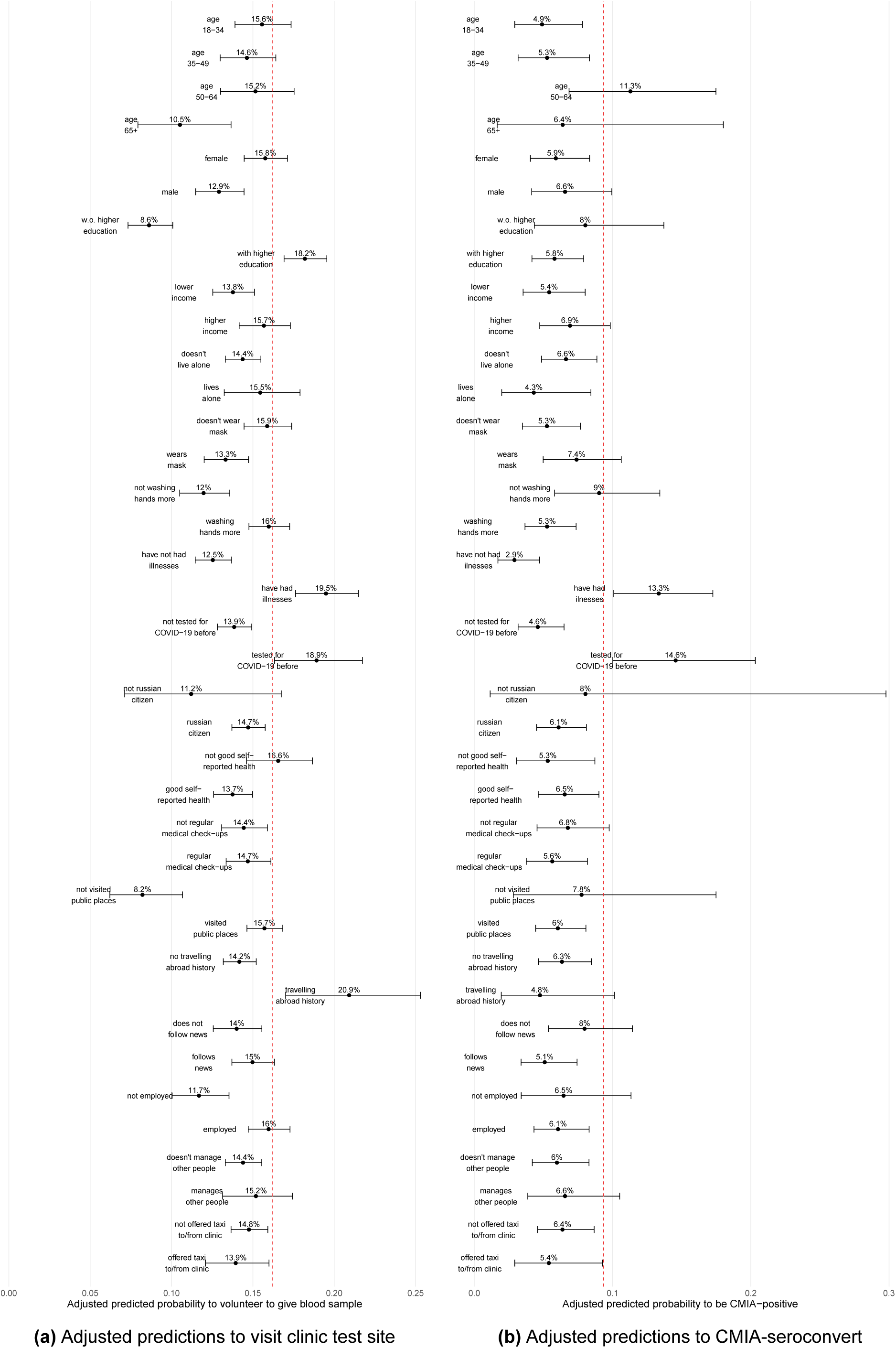
Adjusted predictions to participate in the study or seroconvert. This figure reports adjusted predictions from probit model of individual visiting clinic test site (subfigure a) or being CMIA-seropositive (subfigure b) holding all but one regressor at mean levels. Horizontal lines are 95% CI. Dashed vertical red line is unconditional mean propensity.

